# Determining the source of transmission of SARS-CoV-2 infection in a healthcare worker

**DOI:** 10.1101/2020.04.27.20077016

**Authors:** Nasia Safdar, Gage K. Moreno, Katarina M. Braun, Thomas C. Friedrich, David H. O’Connor

## Abstract

**Background:** Healthcare workers (HCWs) are at the frontlines of the COVID-19 pandemic and are at risk of exposure to SARS-CoV-2 infection from their interactions with patients and in the community (1, 2). Limited availability of recommended personal protective equipment (PPE), in particular N95 respirators, has fueled concerns about whether HCWs are adequately protected from exposure while caring for patients. Understanding the source of SARS-CoV-2 infection in a HCW – the community or the healthcare system – is critical for understanding the effectiveness of hospital infection control and PPE practices. In Dane County, Wisconsin, community prevalence of SARS-CoV-2 is relatively low (cumulative prevalence of ~0.06% – positive cases / total population in Dane county as of April 17). Although SARS-CoV-2 infections in HCWs are often presumed to be acquired during the course of patient care, there are few reports unambiguously identifying the source of acquisition.

**Objective:** To determine the source of transmission of SARS-CoV-2 in a healthcare worker.

## Methods and Findings

Here we report a case of SARS-CoV-2 in a HCW at UW Hospital who performed direct patient care for two non-critically ill patients with confirmed SARS-CoV-2 infections (patient 1 and patient 2). While caring for these patients, the HCW wore a barrier mask, face shield, gowns and gloves. Four days after providing care for these patients, the HCW began experiencing headache, fever, and sore throat. A nasopharyngeal (NP) swab was obtained and tested positive for SARS-CoV-2 viral RNA. To establish the most likely source of infection, we interviewed the HCW’s family member (HCW-F), who had developed a febrile illness eight days before onset of symptoms in the HCW, but was not tested initially due to limited testing availability. We obtained a NP swab from HCW-F which also tested positive for SARS-CoV-2. Figure 1 shows the timeline of events in the four individuals (HCW, HCW-F, Patient 1, and Patient 2) evaluated in this report.

**Figure 1.**
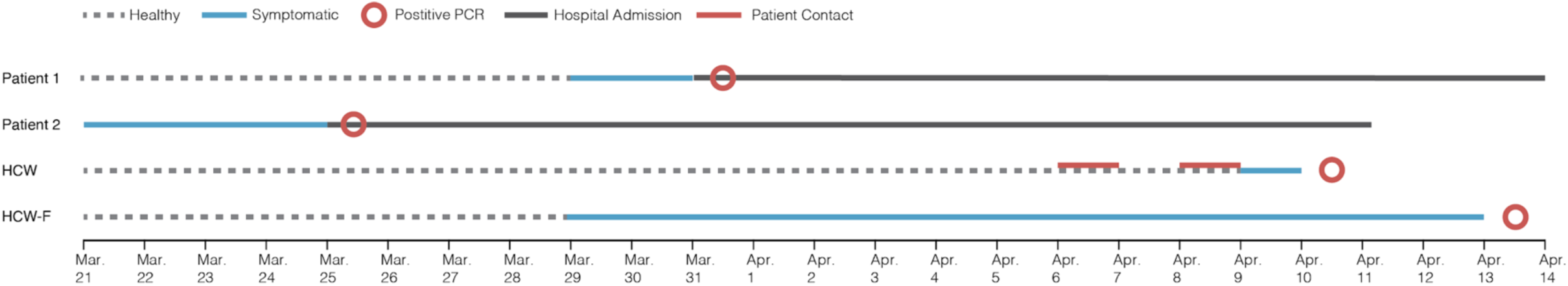
Timeline of infection, contact and testing in the four individuals.

We sequenced viral RNA isolated from NP swab obtained from the two hospitalized patients the HCW treated, the HCW, and the HCW’s family member. We compared consensus SARS-CoV-2 sequences from these four individuals to determine whether the HCW likely acquired infection in a healthcare setting or the community.

Samples from all four individuals were prepared for sequencing using the <u>ARTIC protocol</u> (3) and were sequenced on an Oxford Nanopore GridION device. Consensus sequences were derived using a modified version of the <u>ARTIC bioinformatics protocol</u> (4) that analyzes data once 100,000 reads are obtained from each sample (analysis pipelines are available on <u>GitHub</u>) (5). Figure 2 shows single nucleotide variants (SNVs) that differ at the consensus level between each sample and the original Wuhan reference sequence (Genbank: MN908947.3). The HCW sequence is identical to the HCW-F sequence and distinct from both hospitalized patients (Patient 1 and 2).

**Figure 2.**
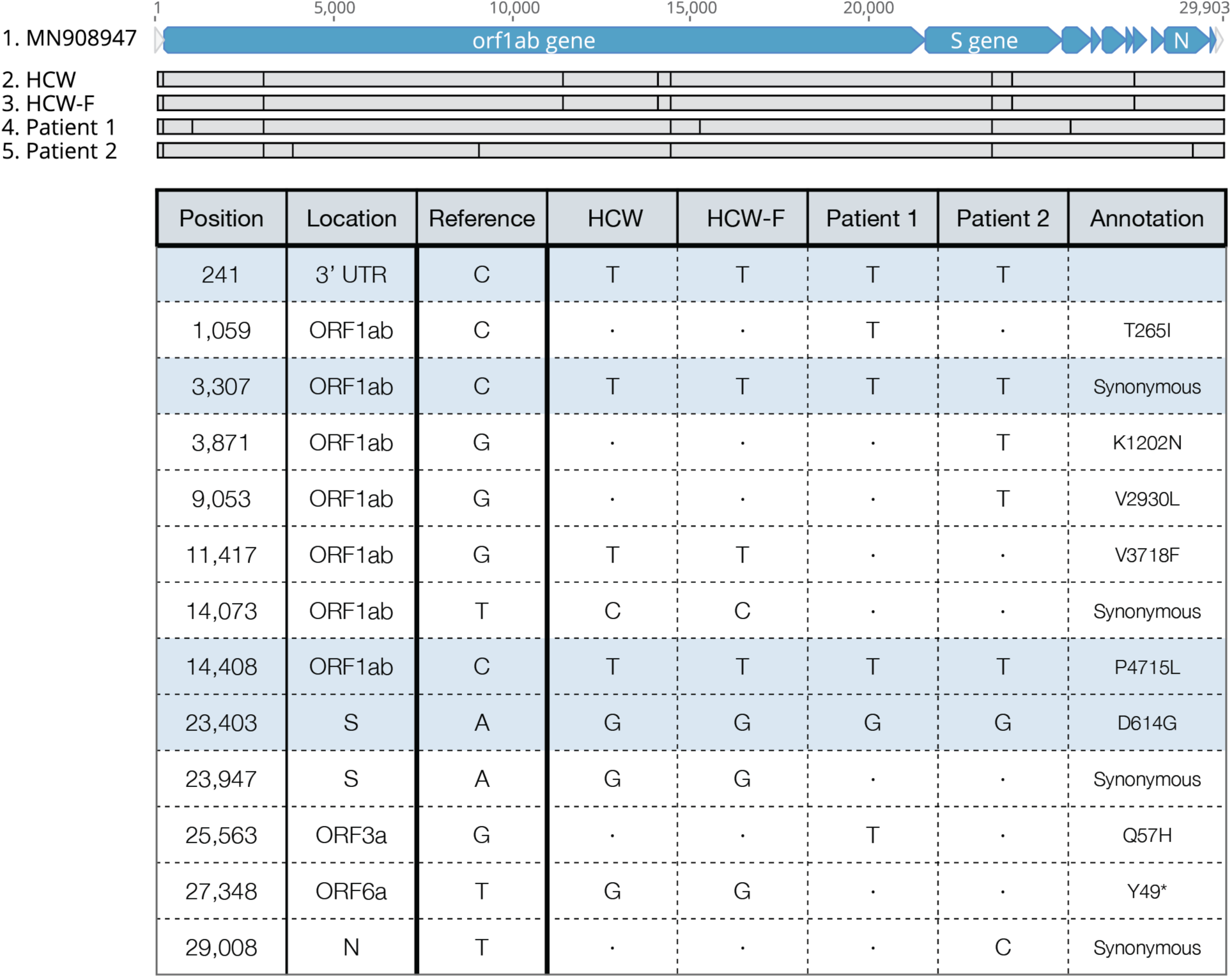
SARS-CoV-2 consensus-level single nucleotide variants. The top alignment image depicts the SARS-CoV-2 genome for all individuals evaluated in this report and highlights SNVs identified relative to the original SARS-CoV-2 Wuhan reference (Genbank: MN908947.3). The table contains additional information about each of these SNVs. A2a clade-defining mutations are shaded light-blue. A dot “.” indicates identity with reference.

## Discussion

While we cannot formally exclude the possibility that the HCW was infected by another asymptomatic, untested hospitalized patient, the identical viral sequences found in the HCW and HCW’s family member provide strong circumstantial evidence for chain of transmission occurring outside of the hospital. Sequencing the HCW’s virus facilitated rapid identification of the likely source of infection – community transmission. This offers reassurance to HCWs currently treating COVID-19 patients that appropriate PPE protects against hospital-acquired SARS-CoV-2 infection. Conversely, demonstrating nosocomial acquisition by sequencing would have provided an impetus for revisiting infection control strategies. Based on these results, viral sequencing of SARS-CoV-2 from HCWs and known contacts both within and outside of patient care settings should be considered an essential part of a comprehensive strategy to protect the health of HCWs and other frontline responders.

## Ethics

The University of Wisconsin-Madison Institutional Review Board deemed this study quality improvement, rather than research, and so declined to subject this study to full review.

## Data Availability

Raw data files are available on GISAID (EPI_ISL_425167, EPI_ISL_425175, EPI_ISL_425176, EPI_ISL_427427) and SRA in BioProject PRJNA614504 (SRX8114920, SRX8114929, SRX8114930). All bioinformatic pipelines used to analyze the data are available on GitHub in the following public repository: https://github.com/katarinabraun/SARS-CoV-2_sequencing/tree/master/Pipelines_to_process_data/Nanopore_pipeline_ARTIC.

## Acknowledgement

We thank Fauzia Osman for creating Figure 1.

